# Circulating inflammatory proteins and risk of Parkinson’s disease and other neurodegenerative disorders: a two-sample Mendelian randomization study

**DOI:** 10.1101/2024.05.03.24306837

**Authors:** Zhichun Chen, Guanglu Li, Liche Zhou, Lina Zhang, Yong You, Jun Liu

## Abstract

**Background:** Accumulating studies have suggested associations between peripheral inflammation and neurodegenerative disorders, including Parkinson’s disease (PD).

**Objective:** To evaluate the causal associations between 91 plasma inflammatory proteins and 4 neurodegenerative disorders.

**Methods:** Two-sample Mendelian randomization studies were performed using summary statistics extracted from genome-wide association studies of 91 plasma inflammatory proteins and 4 neurodegenerative disorders.

**Results:** Genetically proxied tumor necrosis factor receptor superfamily member 9 levels were causally associated with reduced risk of PD (odds ratio [OR] = 0.82, 95% confidence interval [CI] = 0.74-0.92, *p* = 4.18 x 10^−4^, Bonferroni-corrected *p* < 0.05 for 91 proteins). Additionally, we identified potential causal associations between the levels of C-C motif chemokine 20 (OR = 1.14, 95%CI = 1.03-1.25, *p* = 1.29 x 10^−2^) and Alzheimer’s disease, between levels of leukemia inhibitory factor receptor (OR = 0.91, 95%CI = 0.84-0.98, *p* = 1.12 x 10^−2^) and tumor necrosis factor-β (OR = 0.95, 95%CI = 0.93-0.98, *p* = 1.01 x 10^−3^) and amyotrophic lateral sclerosis, between levels of adenosine deaminase (OR = 0.81, 95%CI = 0.71-0.94, *p* = 5.14 x 10^−3^) and interleukin-18 (OR = 0.81, 95%CI = 0.69-0.96, *p* = 1.68 x 10^−2^) and multiple sclerosis.

**Conclusions:** Our study unveils plausible causal associations between circulating inflammatory factors and risk of 4 neurodegenerative disorders. These findings hold promise for promoting risk assessment and prevention of neurodegenerative disorders, meriting further exploration.

## Introduction

Although current medical and health services have dramatically increased the life expectancy of elderly population, the clinical management of neurodegenerative disorders (NDDs) remains a global health challenge ^1-7^. NDDs are characterized by the progressive degeneration of neural cells in central or peripheral nervous system, resulting in the impairment of motor, sensory, cognitive, emotional, and autonomic processes. Alzheimer’s disease (AD), Parkinson’s disease (PD), amyotrophic lateral sclerosis (ALS), and multiple sclerosis (MS) are the major types of NDDs, which significantly reduce the life expectancy and quality of life ^6,7^.

Over the past decade, increasing evidence has shed light on the important role of peripheral inflammation in the pathogenesis of NDDs ^8^. In MS, peripheral immune cells have been shown to trigger neuroinflammation and subsequent demyelination during disease flares ^9, 10^ and blocking of circulating T cell infiltration into the brain parenchyma with natalizumab has been shown to be an alternative targeted therapy in clinical management ^11, 12^. In addition, the mobilization and activation of peripheral immune cells have also been demonstrated to be key drivers of neurodegeneration in AD ^8, 13^, PD ^8, 14^, and ALS ^8, 15, 16^, thus, peripheral inflammation is also an invariant and fundamental feature of AD, PD, and ALS ^8, 17^. However, due to the complex compositions of peripheral immune system ^8, 17^, it remains unknown which molecular and cellular components of peripheral inflammation are causally involved in the pathogenesis of NDDs.

Mendelian randomization (MR) is a powerful method that uses randomly assigned genetic variants as proxies of exposures and infers the causal associations between exposure and outcome ^18, 19^. Considering the critical role of peripheral inflammation in the occurrence of NDDs, it is important to examine whether peripheral immune molecules or cells were causally associated with the risk of NDDs using MR analysis. Previous studies have used MR analysis to estimate the causal associations between immune cell traits and neurological diseases, such as AD^20, 21^, PD^21, 22^, ALS^21, 23^, MS^24, 25^, myasthenia gravis^26^, and subarachnoid hemorrhage^27^. However, how the circulating inflammatory proteins causally associate with NDDs remains to be identified. Zhao *et al*. (2023) conducted a genome-wide protein quantitative trait locus (pQTL) study of 91 circulating inflammatory proteins in 14,824 participants and identified 180 pQTLs (59 *cis*, 121 *trans*) ^28^. In this study, we comprehensively examined the causality between the 4 NDDs and 91 plasma inflammatory proteins using a two-sample MR study.

## Methods

### Ethical approval

This MR study utilized summary statistics from previous published GWAS with ethical approval by corresponding ethics committee, therefore, additional ethical approval for current study is not required.

### Study design

The overall study flowchart is illustrated in **Fig. 1**. MR analyses were performed in accordance with the STROBE-MR checklist ^29^ and Burgess *et al*.’s guidelines ^30^. In this study, we treated the plasma levels of each inflammatory protein as the exposure (a total of 91 inflammatory proteins; sample size: n = 14,824) ^28^, and the risk of each NDD (PD, AD, ALS, and MS; sample size: n = 115,803 ∼ 482,730) as the outcome in the MR analysis ^31-34^. According to the standard protocols of MR study ^18, 19, 29, 30^, each instrumental variable (IV) should satisfy three assumptions as shown below: (i) The IV is significantly associated with the exposure; (ii) The IV is independent of all the other IVs and potential confounding factors; (iii) The IV affects the outcome only through the exposure. During the MR analysis, multiple statistical approaches, such as inverse variance weighted (IVW) method and weighted median method, were performed, however, only IVW was selected to be the major method to examine causal associations due to its high statistical power ^35, 36^. To guarantee the statistical robustness of MR analysis, we conducted comprehensive downstream analyses to evaluate potential biases, which might undermine the reliability of our findings. Specifically, we applied MR-RAPS (Robust adjusted profile score)^37^ to assess the statistical robustness of MR analysis with weak genetic IVs as previously described ^35, 38^. The code of MR-RAPS is available at https://github.com/qingyuanzhao/mr.raps. MR-Egger regression ^39^ and Mendelian Randomization Pleiotropy RESidual Sum and Outlier (MR-PRESSO; https://github.com/rondolab/MR-PRESSO/) ^40^ tests were performed to assess potential bias due to pleiotropy. Leave-one-out (LOO) analyses were conducted to evaluate the bias caused by individual predominant IVs ^35^. The reverse MR analyses, steiger test of directionality, and steiger filtering were performed to exclude the possibility of reverse causation. All statistical analyses were conducted by TwoSampleMR package installed in R software (version 4.3.1; R Foundation for Statistical Computing, Vienna, Austria).

**Fig. 1.**
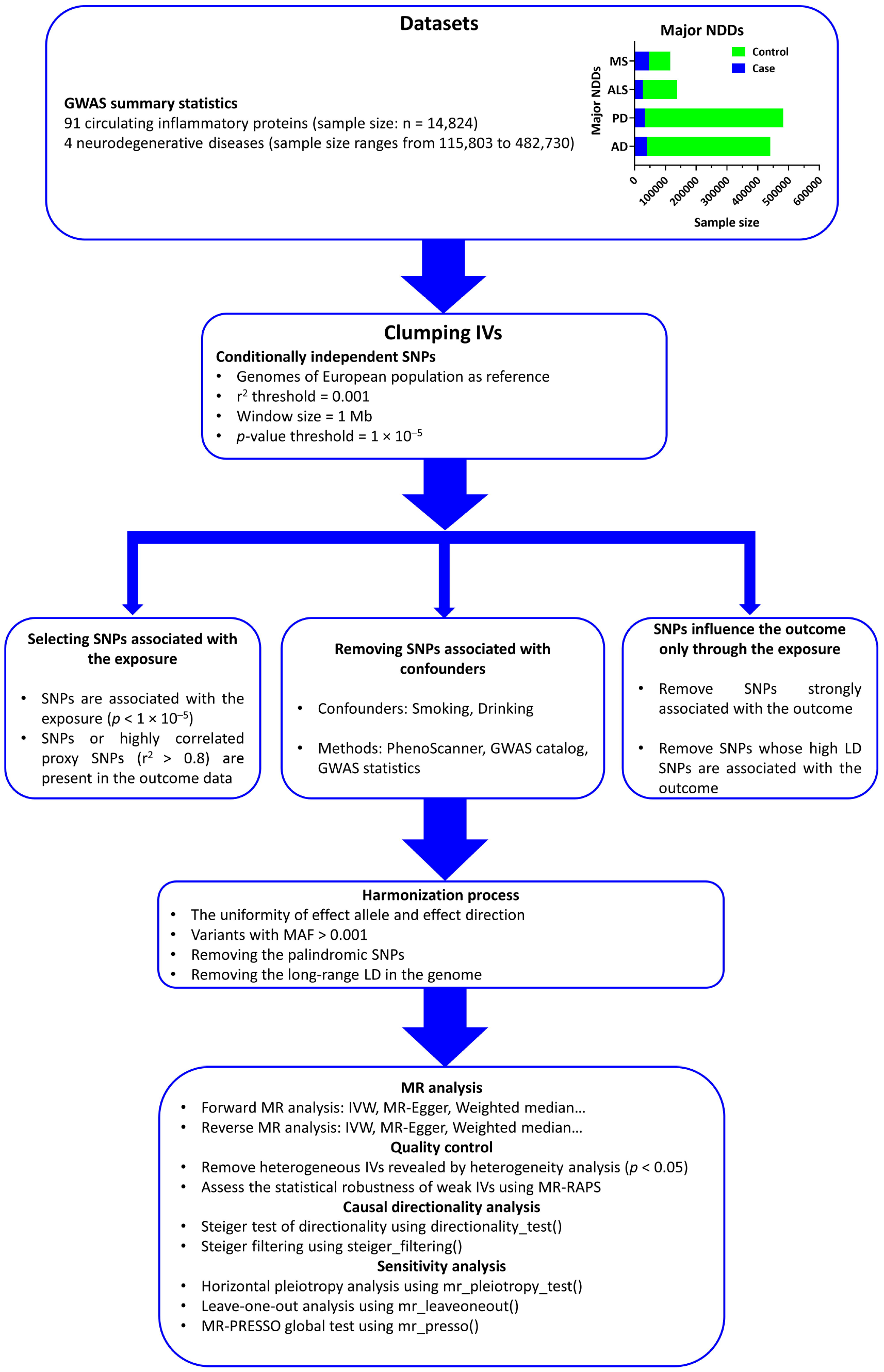
Workflow of the causal inference between circulating inflammatory proteins and NDDs. Abbreviations: NDD, Neurodegenerative disorder; AD, Alzheimer’s disease; PD, Parkinson’s disease; ALS, amyotrophic lateral sclerosis; MS, multiple sclerosis; GWAS, Genome-wide association study; SNP, single nucleotide polymorphism; IV, Instrumental variable; LD, Linkage disequilibrium; MAF, minor allele frequency; MR, Mendelian randomization; IVW, Inverse variance weighted; MR-RAPS, Mendelian randomization-robust adjusted profile score; MR-PRESSO, Mendelian Randomization Pleiotropy RESidual Sum and Outlier.

### Exposure data

GWAS data of circulating inflammatory proteins were sourced from https://www.phpc.cam.ac.uk/ceu/proteins and the EBI GWAS Catalog (accession numbers GCST90274758 to GCST90274848) ^28^. This dataset comprised the genome-wide protein quantitative trait locus (pQTL) mapping of 91 plasma inflammatory proteins measured using the Olink Target platform in 14,824 participants of 11 cohorts ^28^.

### Outcome data

The GWAS summary statistics of PD (GWAS ID: ieu-b-7) ^32^, AD (GWAS ID: ieu-b-2) ^33^, ALS (GWAS ID: ebi-a-GCST90027163) ^34^, and MS (GWAS ID: ieu-b-18) ^31^ were all extracted from IEU OpenGWAS project (https://gwas.mrcieu.ac.uk/). The GWAS of PD involved 33,674 PD cases and 449,056 control individuals from International Parkinson’s Disease Genomics Consortium (IPDGC) ^32^. The GWAS of AD was conducted in 21,982 AD cases and 41,944 controls collected from multiple international cohorts ^33^. The GWAS of ALS was conducted in 27,205 ALS cases and 110,881 controls ^34^. The GWAS of MS was performed in 47,429 MS cases and 68,374 controls from International Multiple Sclerosis Genetics Consortium (IMSGC) ^31^. All participants recruited in above original GWAS were of European ancestry. The GWAS summary statistics of above 4 NDDs have been widely utilized in previous MR studies ^36,41^.

### Selection of genetic IVs

To increase the statistical power of MR analysis, our study initially applied a relaxed statistical *p*-value threshold (*p* < 1 x 10^−5^) to screen the IVs as previously used ^38^. Then, the linkage disequilibrium (LD) clumping was further implemented using 1000 Genomes Project Phase 3 reference panel for the European populations within a 10 MB window to select SNPs that were independently (r^2^ < 0.001) associated with circulating inflammatory proteins. If a particular requested SNP was not present in the outcome GWAS, then highly correlated proxy SNPs (r^2^ > 0.8) were searched as IVs. Heterogeneity test was conducted to detect outliers and improve the accuracy and robustness of IVs and Rucker’s Q’ test for MR-Egger model was performed to evaluate heterogeneity or directional pleiotropy ^42^. Funnel plot was also utilized to assess the heterogeneity of IVs. *p* < 0.05 in the heterogeneity test indicated the necessity of IV adjustments.

### Removing confounders

SNPs associated with potential confounders, such as drinking and smoking behavior, were removed. Specifically, PhenoScanner V2 (http://www.phenoscanner.medschl.cam.ac.uk/) and NHGRI-EBI GWAS catalog (https://www.ebi.ac.uk/gwas/docs/file-downloads/) were used to exclude the SNPs associated with confounders.

### Two-sample MR analyses

An IVW regression method with a fixed-effects model was employed as the primary causal inference. Complementary MR analyses were conducted using the MR Egger, weighted median, simple mode, and weighted mode methods to strengthen the validity of the IVW estimates. MR-RAPS provided a robust inference for MR analysis using many weak IVs ^37^. All these methods were implemented by the functions ‘mr_ivw’, ‘mr_egger_regression’, ‘mr_weighted_median’, ‘mr_raps’, and ‘mr_weighted_mode’ in the TwoSampleMR v0.4.26 R package. Bonferroni approach was utilized to correct for multiple hypothesis testing. Significance was defined as Bonferroni-corrected *p* < 0.05 (uncorrected *p* < 5 x 10^−4^ [0.05/91]), whereas uncorrected *p* < 0.05 that did not meet the Bonferroni-corrected threshold was suggested as potential causal association.

### Sensitivity analysis

First, LOO analysis was performed to examine whether the causal association was particularly driven by a single SNP (*p* < 0.05 was regarded as an outlier). Second, MR-PRESSO global test (https://github.com/rondolab/MR-PRESSO/) was used to detect IVs affected by horizontal pleiotropy (*p* < 0.05). Third, an MR-Egger regression was conducted to examine the potential bias of directional pleiotropy. The intercept in the Egger regression was interpreted as evidence of pleiotropy when the value differed from zero (*p* < 0.05).

## Results

The **Fig. 2** summarized the overall MR causal estimates. Among 91 plasma inflammatory proteins, only the levels of tumor necrosis factor receptor superfamily member 9 (TNFRSF9) were found to be causally associated with reduced risk of PD (OR = 0.82, 95%CI = 0.74-0.92, *p* = 4.18 x 10^−4^, Bonferroni-corrected *p* < 0.05 for 91 proteins). By contrast, other circulating inflammatory proteins failed to reach statistical significance. For AD, we identified a potential causal association between the levels of C-C motif chemokine 20 (CCL20) and risk of AD (OR = 1.14, 95%CI = 1.03-1.25, *p* = 1.29 x 10^−2^). For ALS, the levels of leukemia inhibitory factor receptor (LIFR) (OR = 0.91, 95%CI = 0.84-0.98, *p* = 1.12 x 10^−2^) and tumor necrosis factor-β (TNF-β) (OR = 0.95, 95%CI = 0.93-0.98, *p* = 1.01 x 10^−3^) were revealed to confer risk of ALS. For MS, the levels of adenosine deaminase (OR = 0.81, 95%CI = 0.71-0.94, *p* = 5.14 x 10^−3^) and interleukin-18 (IL-18) (OR = 0.81, 95%CI = 0.69-0.96, *p* = 1.68 x 10^−2^) were causally associated with reduced risk of MS. The LOO analyses and other downstream statistical analyses, such as heterogeneity analysis, pleiotropy test, MR-RAPS, Steiger directionality test, and MR-PRESSO test, all supported the reliability of above findings (**Supplementary Table 1, Supplementary Fig. 1-2**). Zhao *et al*. (2023) identified a potential causal association between circulating CD40 levels and MS (OR = 0.75, 95%CI = 0.70–0.82, *p* = 1.20 × 10^−12^) ^28^, however, in this study, we found the causal association (OR = 0.79 95%CI = 0.73–0.85, *p* = 2.91 x 10^−10^) between plasma CD40 levels and MS was biased by a SNP (rs1883832, *p* = 1.07 x 10^−281^ in outcome GWAS) strongly associated with the risk of MS (**Fig. 3A-C**). When the SNP (rs1883832) was removed from the MR models, no significantly causal association was detected (**Fig. 3D-F**).

**Fig. 2.**
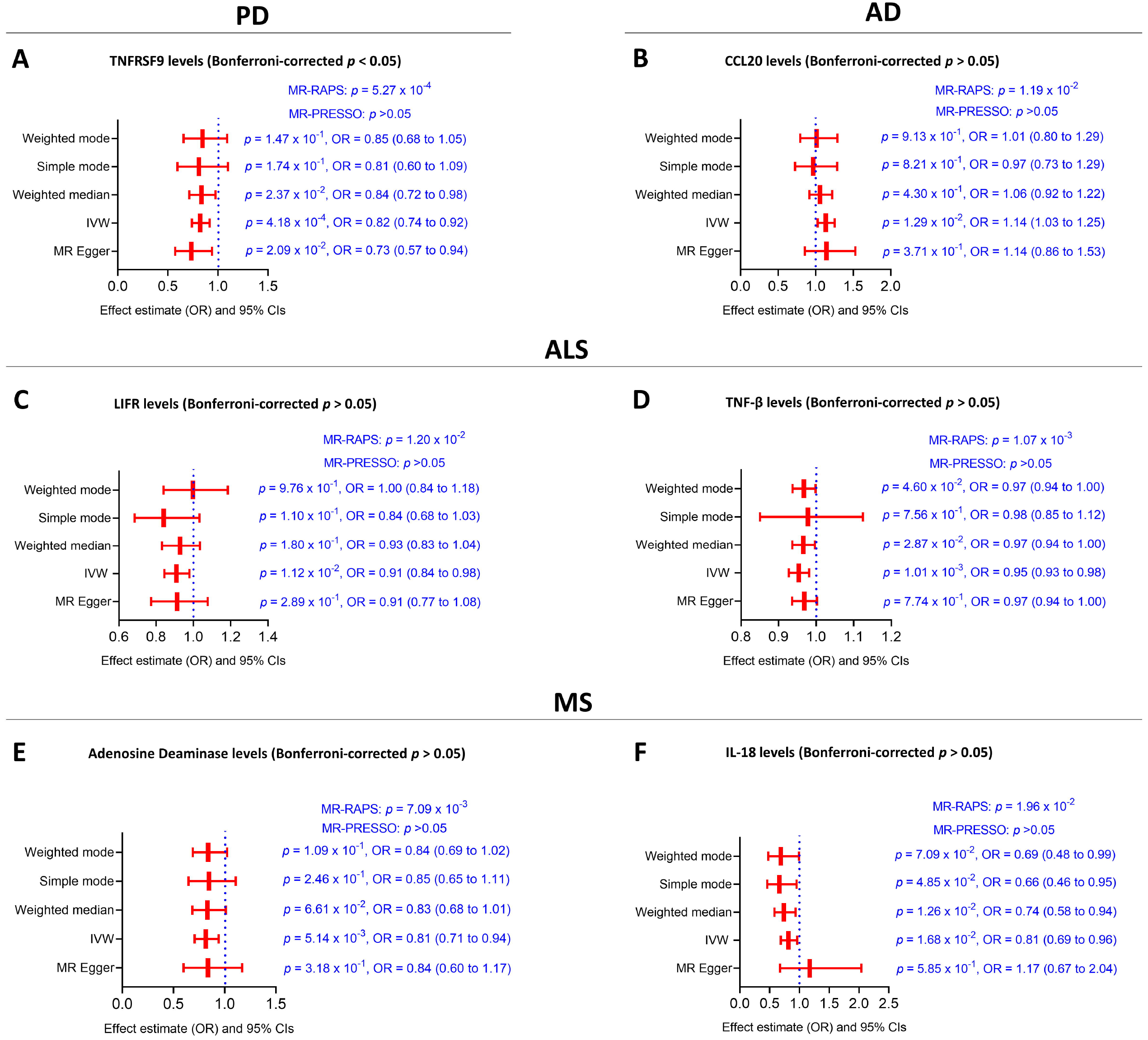
Causalities in the MR analysis. **A-F** The forest plot shows the significant causalities in PD (**A**), AD (**B**), ALS (**C-D**), and MS (**E-F**). The effect estimates represent the OR of disorder and error bars represent 95% CIs. All statistical tests were two sided. *p* < 5 × 10^−4^ after Bonferroni correction was considered significant. Causal effects were estimated using five two-sample MR methods (MR-Egger, IVW, weighted median, weighted mode, and simple mode). Abbreviations: AD, Alzheimer’s disease; PD, Parkinson’s disease; ALS, Amyotrophic lateral sclerosis; MS, Multiple sclerosis; TNFRSF9, Tumor necrosis factor receptor superfamily member 9; CCL20, C-C motif chemokine 20; LIFR, Leukemia inhibitory factor receptor; TNF-β, Tumor necrosis factor-β; IL-18, Interleukin-18; MR, Mendelian randomization; IVW, Inverse variance weighted; MR-RAPS, Mendelian randomization-robust adjusted profile score; MR-PRESSO, Mendelian Randomization Pleiotropy RESidual Sum and Outlier.

**Fig. 3.**
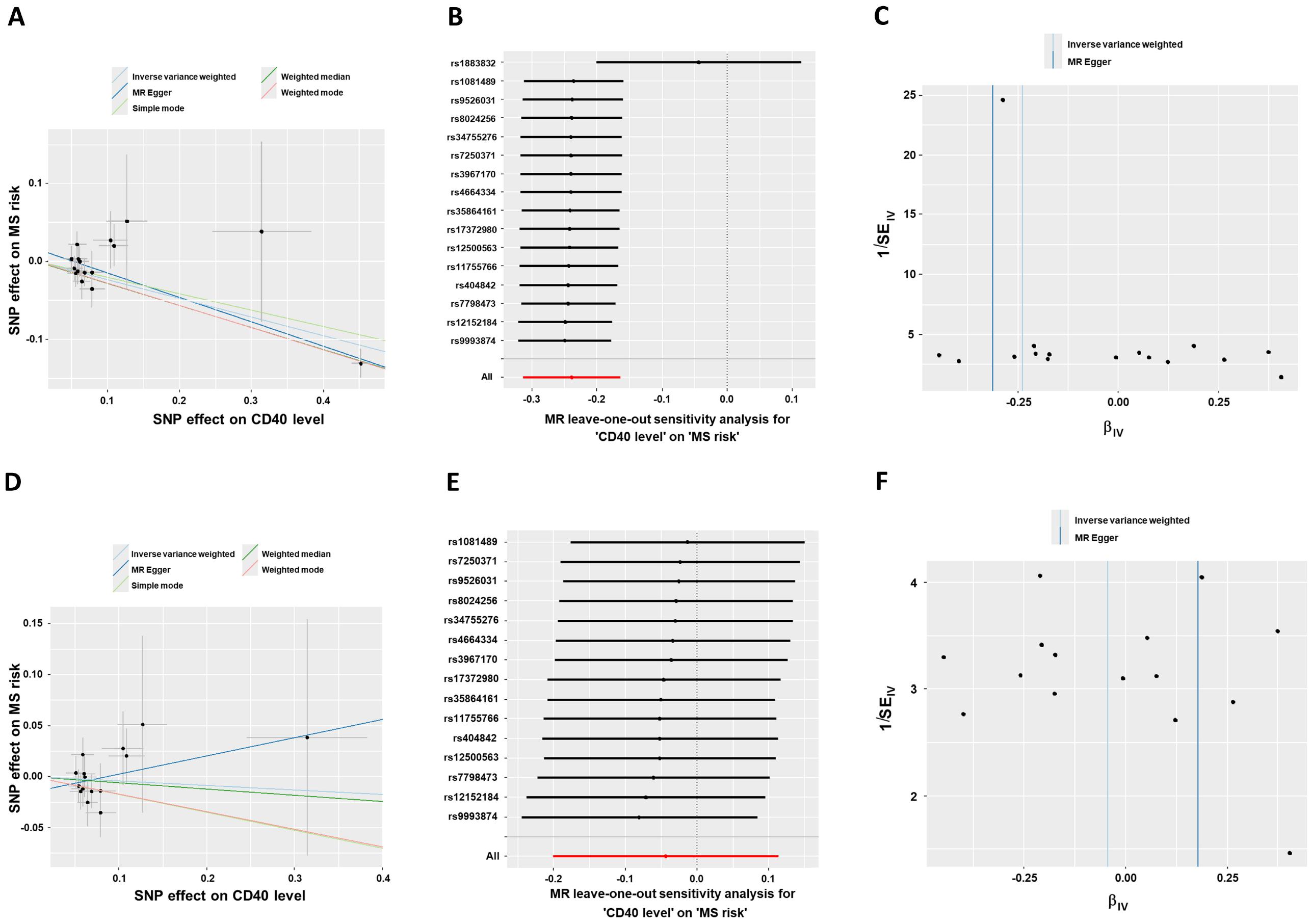
The MR causal estimates between plasma CD40 level and risk of MS. **A-C** The causal association between plasma CD40 levels and MS is determined by rs1883832, which is strongly associated with the risk of MS (*p* = 1.07 x 10^−281^). **D-F** When the rs1883832 was removed from the MR models, no significantly causal association was detected. Abbreviations: SNP, single nucleotide polymorphism; MR, Mendelian randomization; MS, Multiple sclerosis.

## Discussion

Previous evidence has suggested neuroinflammation in central nervous system plays a key role in the pathogenesis of NDDs ^43-45^. Although dysregulated peripheral immunity has also been observed in NDDs according to previous studies^8-10, 15, 16^, whether peripheral immune dysfunction is causally associated with risk of NDDs is largely unknown. In this study, we demonstrate potential causal relationships between several plasma inflammatory proteins and risk of NDDs. Especially, the levels of TNFRSF9 are causally associated with reduced risk of PD by 18%, which even remains statistically significant after Bonferroni correction for multiple hypothesis testing. Our findings support the causality between circulating inflammatory proteins and risk of 4 NDDs, suggesting that peripheral inflammation has a significant clinical influence on 4 NDDs.

The dysregulations of peripheral immunity have been observed in 4 NDDs ^8-10, 15-17^. For innate immunity, both monocytes and macrophages have been revealed to be involved in the occurrences of NDDs, including PD ^46-48^, ALS ^49, 50^, and MS ^51, 52^. Abnormal activations of monocytes have been observed in PD patients ^47, 48^ and monocytes seem to exhibit age-dependent impairment of α-synuclein oligomer uptake ^53^. In ALS, the modification of peripheral macrophages could suppress proinflammatory microglial responses and exert neuroprotection ^50^. In MS, remarkable bone marrow myelopoiesis has been shown to increase the invasion of peripheral neutrophils and Ly6C^high^ monocytes into brain parenchyma ^52^. Especially, Cxcl10^+^ monocytes have been shown to induce experimentally autoimmune encephalomyelitis, a model of MS ^54^. As to adaptive immunity, infiltration of peripheral CD8+ T cells into AD cultures could induce abnormal microglial activation, neuroinflammation and neurodegeneration ^13^. Additionally, PD patients exhibit reduced blood CD8+ cytotoxic T cells, which are primarily associated with the severity of the disease. Importantly, peripheral Th17 lymphocytes from PD patients have been demonstrated to induce neuronal cell death in an iPSC-based PD model ^55^. In ALS, increased blood CD4+EOMES+ T cells are associated with poor prognosis ^56^ and T cells have been shown to infiltrate into the spinal cord of sporadic ALS patients ^57^. In MS, blood memory B cells are demonstrated to activate autoreactive CD4+ T cells to trigger neuroinflammation and demyelination ^58^. All these findings suggest that peripheral immune dysregulation is a remarkable pathological marker of NDDs. With MR analysis, previous studies have also shown several blood immune cell traits are causally correlated with 4 NDDs, including AD^20, 21^, PD^21, 22, 59^, ALS^21, 23^, and MS^24, 25^. In this study, we provided new evidence that peripheral inflammatory proteins are also causally associated with NDDs, which further support the causal role of peripheral immune dysregulation in the pathogenesis of NDDs. TNFRSF9 is a receptor for TNFSF9/4-1BBL (also known as CD137L), which is a type II membrane protein of the TNF superfamily. TNFRSF9 could enhance the survival, cytotoxicity, and mitochondrial activity of CD8+ T cell, thereby enhancing immunity against viruses and tumors ^60, 61^. The levels of TNFRSF9 are increased in PD patients ^62^, especially in those with cognitive impairment ^62^. Patients with mutations in *DJ-1* develop early-onset PD and exhibit slow disease progression, while a novel variant that contains the partial deletion of neighboring genes *DJ-1* (del exons 1-5) and *TNFRSF9* (del exons 1-6) is revealed to cause a new type of juvenile PD with remarkably earlier disease onset ^63^. In current study, we find levels of TNFRSF9 are associated with reduced risk of PD, which indicates TNFRSF9 may play a protective role in PD patients. However, according to previous literature, the specific mechanisms of TNFRSF9 involved in PD pathophysiology have not been investigated before. The dysregulations of T cell immunity have been shown to induce neuroinflammation and dopaminergic neurodegeneration in PD patients ^64-67^. Considering the key role of TNFRSF9 in the modulation of CD8+ T cells ^60, 61^, we hypothesize that TNFRSF9 may regulate CD8+ T cells to participate in the neuroinflammation of PD patients. Future studies are required to further decode the molecular and cellular mechanisms underlying the effects of TNFRSF9 in PD patients. We find the levels of CCL-20 are associated with increased risk of AD, which is consistent with previous studies showing that chemokines are associated with cognitive impairment in AD patients ^68^. Although the specific role of CCL-20 in AD has not been studied before, other chemokines, such as CXCL1 and CXCL10, have been shown to mediate neuroinflammation and neurodegeneration in AD models ^13, 69^. LIFR is a receptor for LIF, which is a member of the IL-6 cytokine family. *LIF* gene has been shown to be modifier gene in the pathogenesis of ALS ^70^. Interestingly, a recent MR analysis has also revealed that LIFR levels are causally associated with ALS risk ^71^, which are consistent with our results. Adenosine deaminase is a key enzyme in purine salvage pathways and the mutations in adenosine deaminase gene cause autosomal recessive severe combined immunodeficiency (SCID). MS patients exhibit elevated levels of adenosine deaminase in CSF ^72^ and elevated adenosine deaminase activities in serum ^73^. Besides, the distribution of adenosine deaminase isoenzymes has been shown to be impaired in CSF and plasma of MS patients ^74^. It seems that adenosine deaminase may regulate the pathogenesis of MS through adenosinergic signaling ^75^. *IL-18* 607C/A gene promoter polymorphism has been shown to be a major genetic factor for MS ^76^ and the serum levels of IL-18 are increased in MS patients ^77, 78^. In this study, we found the plasma IL-18 levels are associated with reduced risk MS patients, indicating IL-18 may play an essential role in the occurrence of MS.

Previous MR analysis reported a causal association between plasma CD40 levels and MS risk ^28^, however, we didn’t completely duplicate their results. In their analysis, they used quasi-independent variants (r^2^ < 0.1) to select IVs, which may increase LD of IVs and bias of their results. Actually, we found the causal association between plasma CD40 levels and MS may be biased by a risk variant of MS, rs1883832 ^31^. rs1883832 is a *cis*-pQTL of plasma CD40 and the GTEx project has shown that rs1883832 C allele is associated with increased CD40 levels in both blood (*p* < 1.6 x 10^−13^) and cerebral cortex (*p* < 5.6 x 10^−7^). Additionally, previous studies have shown that the CD40–CD40L costimulatory pathway is implicated in the pathogenesis of MS ^79-81^ and anti-CD40L monoclonal antibody frexalimab has been shown to reduce the number of new gadolinium-enhancing T1-weighted lesions in MS patients ^82^. Based on these results, we conclude that CD40–CD40L pathway may be causally associated with pathogenesis of MS, however, whether plasma CD40 levels are causally associated with MS risk remains to be further elucidated.

In conclusion, our study indicates the peripheral inflammation may be causally associated with 4 NDDs. Future studies are encouraged to decipher the molecular mechanisms underlying the associations between circulating inflammatory proteins identified in this study and the pathogenesis of 4 NDDs.

## Data Availability

The data supporting the findings of this study are available from the corresponding author upon reasonable request.

## Author contributions

ZC had full access to all of the data in the study and takes responsibility for the integrity of the data and the accuracy of the data analysis. Concept and design: ZC, YY, and JL. Acquisition, analysis, or interpretation of data: ZC, GL, LZ, and LZ. Drafting of the manuscript: ZC and JL. Critical revision of the manuscript for important intellectual content: ZC, LZ, YY, and JL. Statistical analysis: ZC and GL. Obtained funding: YY and JL.

## Supplementary information

Supplementary data related to this article can be found online.

