## Supplementary materials for "Circulating inflammatory proteins and risk of Parkinson’s disease and other neurodegenerative disorders: a two-sample Mendelian randomization study"

**Supplementary Table 1. Statistics used to assess the reliability of MR analysis**

| NDDs/Exposure  Statistical analysis | PD | AD | ALS | | MS | |
| --- | --- | --- | --- | --- | --- | --- |
|  | TNFRSF9 level | CCL20 level | LIFR level | TNF-β level | Adenosine deaminase level | IL-18 level |
| Heterogeneity analysis | *p* = 0.68 | *p* = 0.36 | *p* = 0.49 | *p* = 0.89 | *p* = 0.95 | *p* = 0.58 |
| Pleiotropy test | *p* = 0.32 | *p* = 0.95 | *p* = 0.96 | *p* = 0.13 | *p* = 0.87 | *p* = 0.21 |
| MR-PRESSO | *p* = 0.71 | *p* = 0.44 | *p* = 0.54 | *p* = 0.67 | *p* = 0.97 | *p* = 0.51 |
| MR-RAPS | *p* = 5.27 x 10^-4^ | *p* = 0.01 | *p* = 0.01 | *p* = 0.001 | *p* = 0.01 | *p* = 0.02 |
| Steiger directionality test | *p* = 1.17 x 10^-164^ | *p* = 4.71 x 10^-93^ | *p* = 5.05 x 10^-125^ | *p* = 0.00 x 10^0^ | *p* = 1.78 x 10^-86^ | *p* = 1.30 x 10^-45^ |
| Steiger filtering | *p* < 0.05 for all | *p* < 0.05 for all | *p* < 0.05 for all | *p* < 0.05 for all | *p* < 0.05 for all | *p* < 0.05 for all |
| Reverse MR (IVW method) | *p* = 0.93 | *p* = 0.48 | *p* = 0.92 | *p* =0.53 | *p* =0.25 | *p* =0.91 |
| Leave-one-out analysis | *p* < 0.05 for all | *p* < 0.05 for all | *p* < 0.05 for all | *p* < 0.05 for all | *p* < 0.05 for all | *p* < 0.05 for all |

**Abbreviations:** NDD, Neurodegenerative disorder; PD, Parkinson’s disease; AD, Alzheimer’s disease; ALS, Amyotrophic lateral sclerosis; MS, Multiple sclerosis; TNFRSF9, Tumor necrosis factor receptor superfamily member 9; CCL20, C-C motif chemokine 20; LIFR, Leukemia inhibitory factor receptor; TNF-β, Tumor necrosis factor-β; IL-18, Interleukin-18; MR, Mendelian randomization; PRESSO, Pleiotropy RESidual Sum and Outlier; RAPS, Robust adjusted profile score; IVW, Inverse-variance weighted.

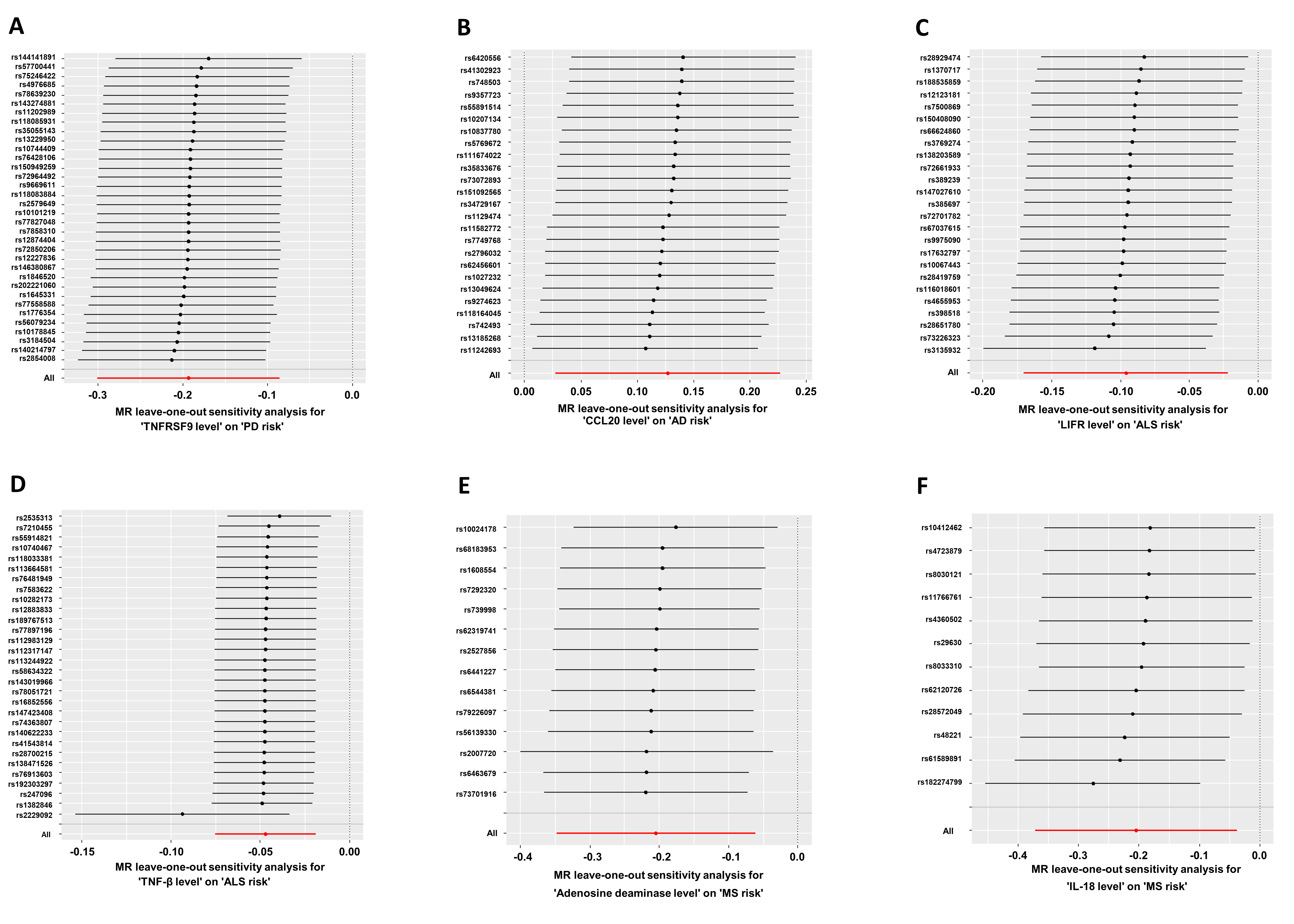

**Supplementary Fig. 1** The LOO analysis for significant MR causal estimates. **A-F** The LOO analysis for significant MR causal estimates in PD (**A**), AD (**B**), ALS (**C-D**), and MS (**E-F**). Abbreviations: AD, Alzheimer’s disease; PD, Parkinson’s disease; ALS, Amyotrophic lateral sclerosis; MS, Multiple sclerosis; TNFRSF9, Tumor necrosis factor receptor superfamily member 9; CCL20, C-C motif chemokine 20; LIFR, Leukemia inhibitory factor receptor; TNF-β, Tumor necrosis factor-β; IL-18, Interleukin-18; MR, Mendelian randomization; LOO, Leave-one-out.

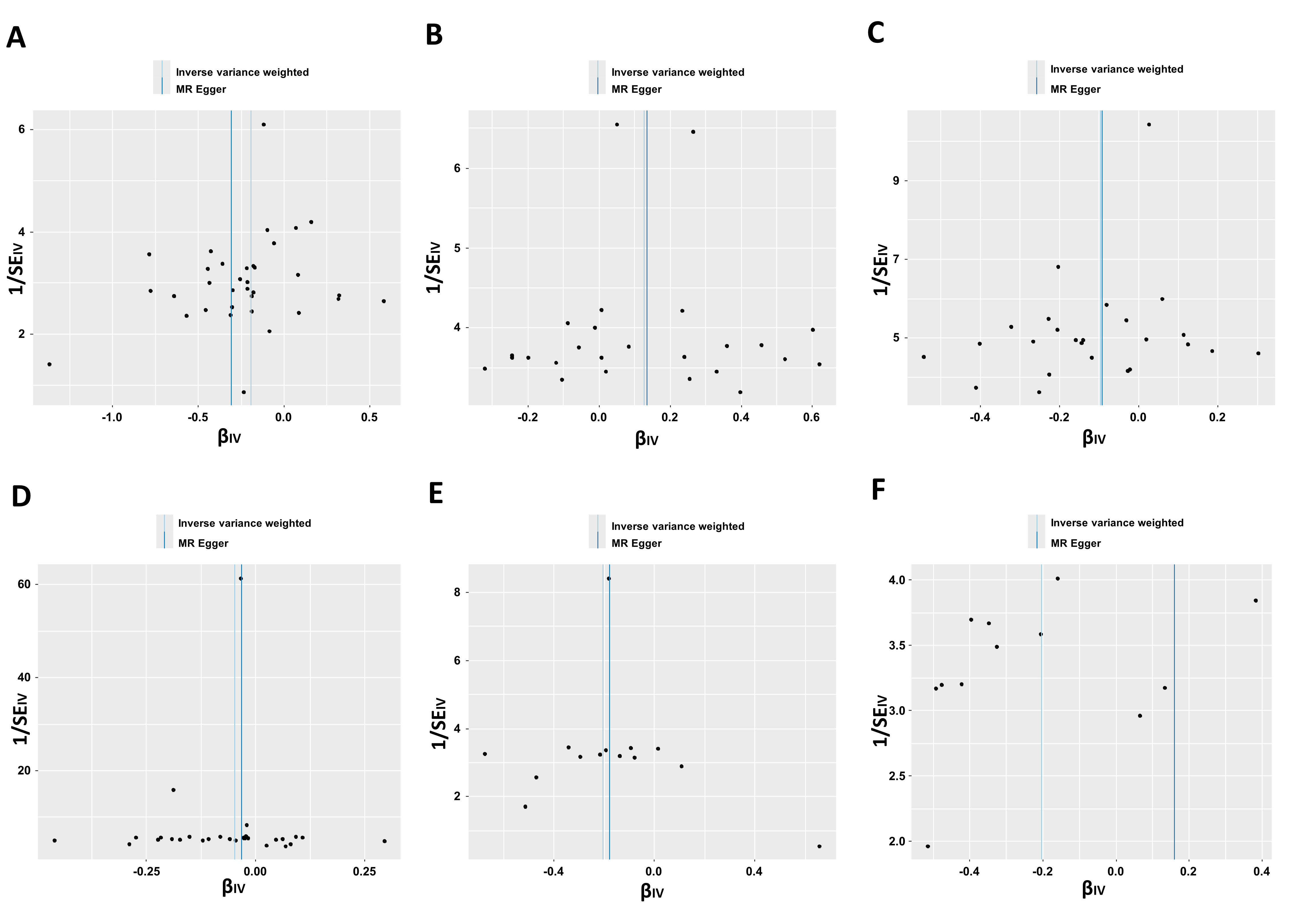

**Supplementary Fig. 2** The funnel plot for significant MR causal estimates. **A-F** The funnel plot for MR results in PD (**A**), AD (**B**), ALS (**C-D**), and MS (**E-F**). Abbreviations: MR, Mendelian randomization.
